# Rapid Competency in NIR-Based Neuroimaging: Training Non-Experts for Head Trauma Triage

**DOI:** 10.64898/2025.12.15.25342308

**Authors:** Sebastian D’Amario, Melissa T. Lamanna, Jason D. Riley, Douglas J. Cook

## Abstract

**Objective:** To evaluate whether novice, non-specialist operators can rapidly learn to use a handheld near-infrared (NIR) head scanner and maintain scan quality after brief training, supporting its use for point-of-care triage when computed tomography is unavailable or delayed.

**Methods:** Thirty-two right-handed adults with no prior NIR experience received a brief standardized training session (∼2 minutes) on the ArcheOptix NIRD® device, which detects intracranial hemorrhage by tracking hemoglobin absorption during guided scalp scans. Operators then completed two full-head scans on a healthy volunteer: an initial competency assessment (Scan 1) and a follow-up assessment after one day without refresher training (Scan 2). Performance metrics included total scan time, frequency of repeat scans prompted by loss of contact or light leaks, and mean scanpath time as an index of handling efficiency and consistency. Scan quality was evaluated using Lift on dark and Noise on dark indices. User experience was measured after each scan with the 10-item System Usability Scale (SUS, 0–100). Within-participant changes were analyzed with paired t tests or Wilcoxon signed-rank tests.

**Results:** All operators completed both sessions. Median performance improved from Scan 1 to Scan 2, with total scan time decreasing from 5 min 27 s to 2 min 53 s. The proportion of “Excellent” scans (<5 minutes) increased from 50% to 84%, and “Poor” scans (>10 minutes) fell to zero. Repeat scans per session declined from 38 to 24, and mean scanpath time shortened while becoming more consistent. Lift on dark and Noise on dark remained stable, indicating no degradation in signal quality as operators worked faster. SUS scores improved from 69.4 to 76.5, reflecting higher perceived ease of use and confidence.

**Conclusions:** After minimal training, novice operators achieved rapid, reliable NIR scans with faster performance, fewer repeat scans, stable signal quality, and improved usability ratings. This work shows that portable NIR can practically complement CT by helping prioritize transport and focus scarce imaging resources in emergency, sideline, and remote head trauma triage.

## Introduction

Traumatic head injuries represent a critical global health burden, with an estimated 69 million individuals affected annually worldwide.^1^ Timely detection of intracranial abnormalities, such as hematomas or edema, is essential to prevent secondary injuries and improve patient outcomes.^2^ While computed tomography (CT) remains the standard for diagnosis,^3^ its reliance on radiation, cost, and limited accessibility in pre-hospital or low-resource settings underscores the need for rapid, portable screening tools. Near-infrared (NIR) devices have emerged as promising alternatives, leveraging the differential absorption of infrared light by hemoglobin to detect intracranial bleeding or oxygenation changes non-invasively.^4–8^ However, clinical utility hinges on technical accuracy and on whether frontline health care providers, many without specialized training, can operate these tools efficiently and reliably.

Near-infrared devices use hemoglobin’s absorption of near-infrared light to identify intracranial hemorrhage after head injury.^9^ This capability enables point-of-care triage in remote environments, particularly when CT scans are unavailable,^6,9–11^ offering a rapid, portable, and effective diagnostic alternative in time constrained emergency settings.^12^ One such device, the NIRD^®^ scanner from ArcheOptix Biomedical, is a handheld unit that scans the scalp to identify acute subdural and epidural hemorrhage.^13^ It uses a single LED at 805nm, has a built-in shield to exclude ambient light and shows a simple, location-marked report on a tablet. As new technologies enter this space, rigorous validation is needed to establish decision thresholds in their operationalization.

Despite advances in NIR device design, scant attention has been paid to the human factors governing their real-world implementation. For these tools to be adopted in emergency departments, sports fields, battlefield settings, and remote environments, they must be usable by personnel with minimal technical expertise after brief training.^8,10,11^ Because adoption depends on trust, standardized protocols should evaluate how quickly novices reach proficiency in NIR scanning and how well operators maintain consistent data quality. This study addresses that need through a structured training program that prepares non-specialist users to perform rapid NIR scans. We focus on four core pillars: (1) rapid learning, assessed by time to competency and repeat scans needed; (2) scan quality, evaluated with signal-integrity; (3) short-term skill retention after a single session, tested with an unassisted repeat scan; and (4) operator confidence and perceived usefulness, captured with a brief questionnaire. Our protocol aims to bridge the gap between technological innovation and practical clinical adoption.

Our findings have implications for the integration of NIR devices into acute care workflows, particularly in settings where rapid triage is paramount. We aim to demonstrate that with a simple and robust designed training protocol, operators with no prior NIR experience can achieve proficiency in ∼5 minutes, producing scans comparable in quality to those of expert users. This advances the field of point-of-care neuroimaging while providing a replicable framework for evaluating the usability of emerging medical technologies across diverse operator populations.

## Methods

### Participants

A cohort of 32 novice right-handed operators with no prior NIR experience was recruited (mean age: 24 ± 3.3, 19 female). All scans were performed on a volunteer with no history of head injury. The expert scanner in comparison is our last author who performed one full scan on the same volunteer. For a within participant comparison, this sample size provides about 80% power at α = 0.05 to detect at least a medium pre to post improvement (Cohen’s *d*_*z*_ ≈ 0.5) in usability outcomes. Ethical approval was obtained from the Queen’s University Health Sciences Research Ethics Board (HSREB#6019089), and all participants provided written informed consent. We excluded participants who have prior experience with facilitating hands-on neurological assessment.

### Training Protocol

We began with a brief conceptual overview regarding the device’s use of light emission and hemoglobin absorption for the detection of bleeding (Figure 1A). This was followed by a quick demonstration of how to use the device, starting with calibration of the device followed by this instruction: “Hold the button down, and with normal pressure, drag the device along the head. You will follow the path lit up on the screen in front of you and will advance to the following path when it lights up to make your way across the whole head” (Figure 1B). They were also informed that if the device requested a rescan, you would need to ensure contact of the device to the head and repeat the scan path (Figure 1B). This entire practice component took around 2 minutes and was standardized for all participants. Participants then performed their competency assessment. They received minimal feedback only reminding them of maintaining contact throughout dragging the device along the path (which the rescan already indicated to them). Participants came back after with 1 full day gap in-between for their second assessment. For this attempt, they received no training or reminder before beginning the competency test.

**Figure 1.**
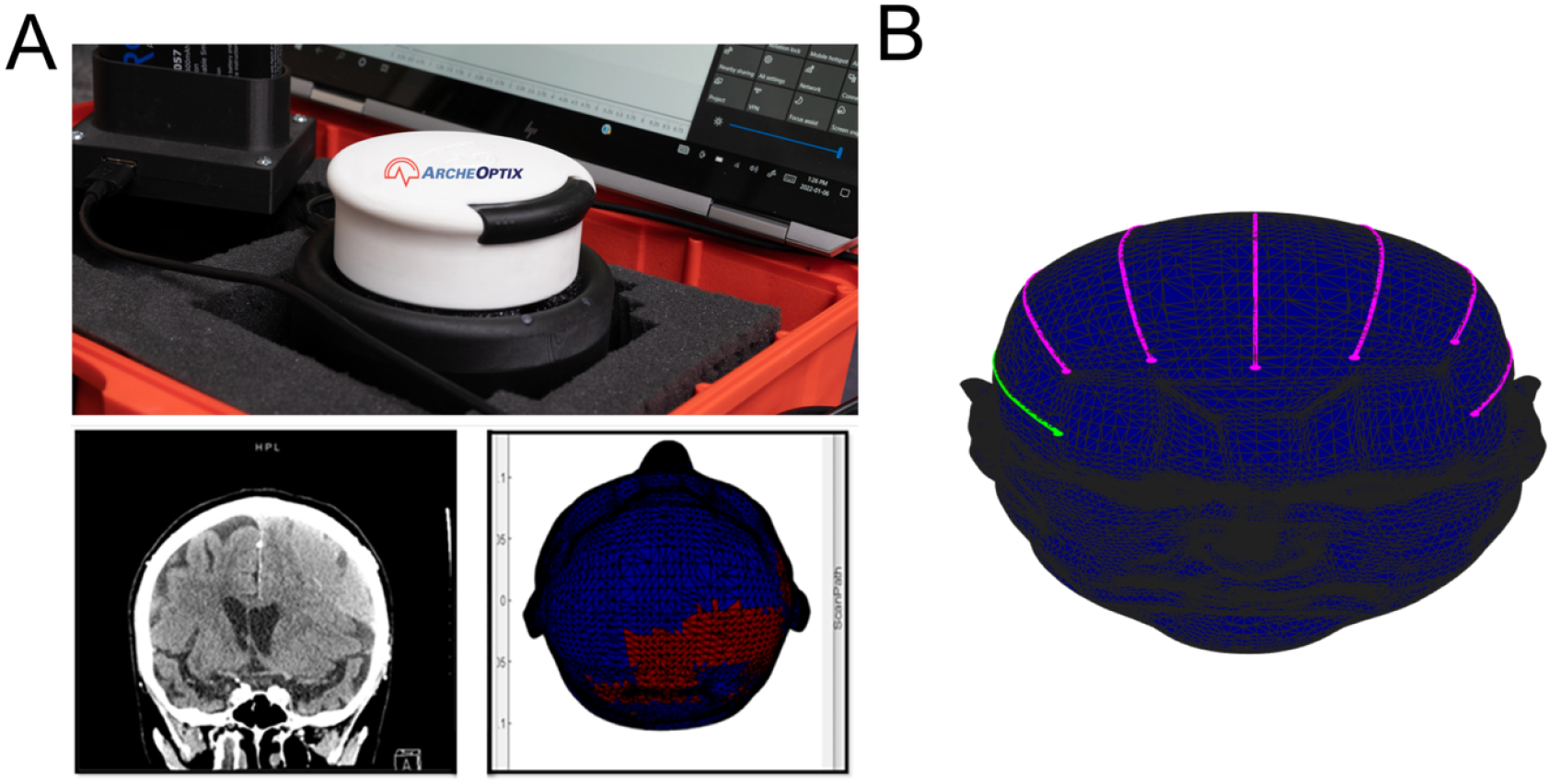
Device overview and guided scanning workflow. **(A)** Conceptual overview of the handheld NIR device (top) that emits light and detects hemoglobin absorption to flag intracranial bleeding (bottom right). **(B)** Guided scan procedure after calibration: operators press and hold the button and move the device with normal pressure along the illuminated path on the display, advancing as each new path lights up to cover the head. If a rescan indicator appears, users re-establish contact and repeat that path before proceeding.

### Data Collection

During data collection, the NIRD^®^ scanner emits near-infrared light while the handheld probe is moved over the scalp and records the diffusely reflected signal at two “near” detectors that sample superficial tissues and two “far” detectors that sample deeper layers.^13^ To separate deep from shallow effects, the device computes a unitless ratio:

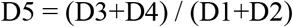

Where D1 and D2 are near-detector intensities and D3 and D4 are far-detector intensities. Deep absorbers such as acute blood reduce the far signal disproportionately, so D5 deviates from its usual range as the probe passes over a hemorrhage. The probe is swept in systematic passes; each D5 value is time-registered to position, then smoothed and interpolated to produce a continuous head map that highlights focal abnormalities. The device obtains a per individual baseline D5 value based on a participant’s skin tone and applies an adaptive threshold using a neighbouring method. This meaning that if an anomaly D5 is detected i.e. a possible bleed, the device checks the D5 directly surrounding that area, and if both show up as inconsistent from baseline, then it will be displayed as a bleed. It is robust in establishing spatial consistency across adjacent passes to suppress artefacts from brief contact loss or hair.^14^ The output is a simple head diagram with a localized highlight where D5 indicates abnormally high absorption, providing an intuitive, real-time screen for intracranial hemorrhage.

Data collection encompassed multiple dimensions of performance, scan quality, and user feedback. We investigated whether operators learned the device quickly, acquired clear scans, retained skills after a single session, and reported confidence in the tool’s usefulness. Performance metrics were captured by recording the speed of each scan, defined as the elapsed time from the beginning of the first scan path to scan completion, and by quantifying the number of rescans throughout the scan. We computed mean scanpath time as the average time to complete a pass along the scan path which informed us how comfortable the user is running the device, and the within-session standard deviation as a measure of consistency. Speed was also categorized as Excellent (<5 min), Moderate (5-10 min), or Poor (>10 min). Scan quality was assessed using Lift on dark, reflecting probe contact during motion, and Noise on dark, reflecting overall clean scan quality, with lower values indicating better performance for both. Usability and perceived value were measured after each scan using the 10-item System Usability Scale (SUS) scored 0-100, capturing ease of use, learnability, confidence, and perceived usefulness.^15^

### Statistical Analysis

We analyzed within-participant change for each outcome. We assessed normality of paired differences with Q-Q plots and the Shapiro-Wilk test. If differences were approximately normal, we used a paired t-test and reported the mean difference, 95% CI, and Cohen’s *d*_*z*_ (effect size for paired designs, computed as the mean of the difference scores divided by their SD). If differences were non-normal or outlier-prone, we used the Wilcoxon signed-rank test and reported the rank-biserial correlation *r*_*rb*_ as the effect size. Significance was set at p < 0.05. Data were analyzed using R v4.5.1.^16^

## Results

All 32 novice operators completed the structured training program. Due to the need for rapid decision making in TBI triage, we predefined a simple rubric based on total scan time: Excellent < 5 minutes, Good 5-10 minutes, Poor > 10 minutes (Figure 2A). From baseline to the follow-up, total scan time shortened 2m 24s on average (Figure 2B) and the share of Excellent runs increased substantially (proportion scan 1 → scan 2: Excellent 50% → 84%; Good 28% → 16%; Poor 22% → 0%; Figure 2A). Across sessions, users became faster or were already fast. Baseline assessments revealed an average scan time of 5m 27s (SD ± 4m 39s; Table 1; Figure 2B) and an average rescan rate of 38 rescans per scan (SD ± 27; Table 1; Figure 2C). Post-training, participants achieved proficiency with a significant reduction in scan time to 2m 53s (SD ± 1m12s; p < 0.001; *r*_*rb*_ *=* -0.69; Table 1; Figure 2B) and rescan rates to 24 rescan per scan (SD ± 11; p < 0.001; *r*_*rb*_ *=* -0.62; Table 1; Figure 2C). These values reduced and became similar to an expert handler of the NIRD^®^.

**Figure 2.**
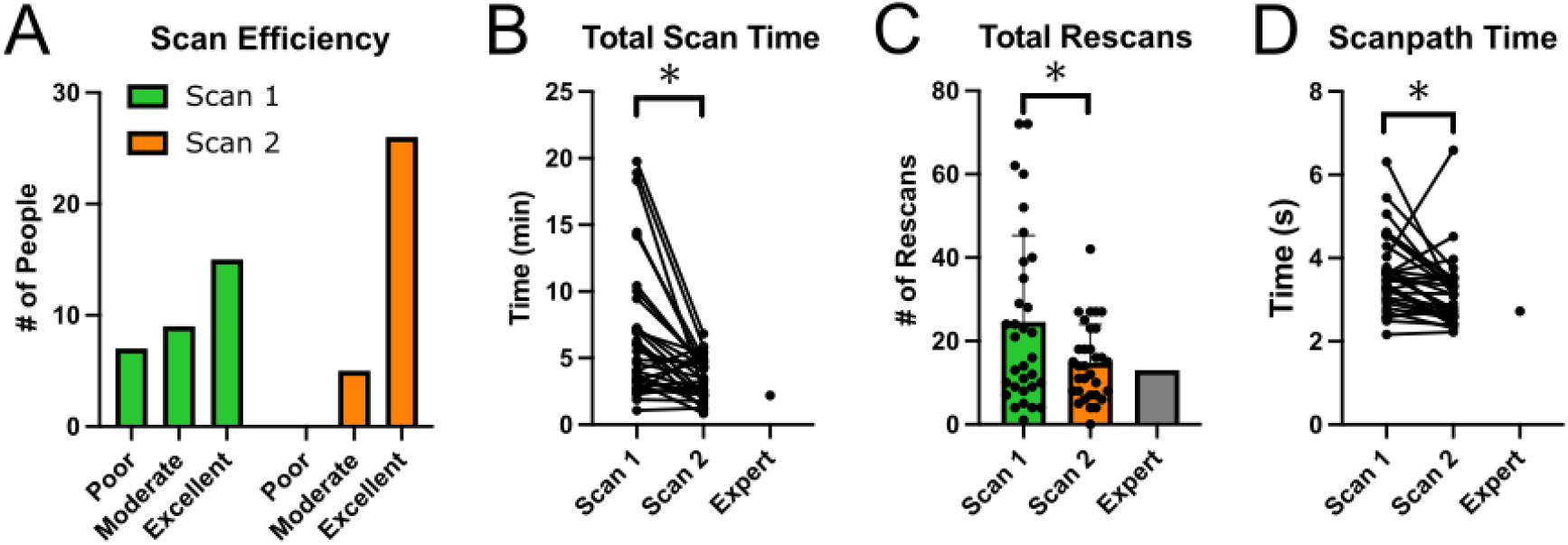
Operators become faster and more consistent with brief practice nearing expert level performance. **(A)** distribution of runs by a simple efficiency rubric based on total scan time (Excellent <5 min, Good 5-10 min, Poor >10 min) for Scan 1 (green) and Scan 2 (orange); **(B)** paired dots of each participant’s total scan time with the expert plotted at right; **(C)** rescans per scan with individual participants overlaid and an expert reference for context; and **(D)** paired dots of mean scanpath time, defined as the average duration of the seven scan paths per session. Across panels, participants shift toward shorter scan times, fewer rescans, and more efficient handling after brief familiarization.

Operator efficiency was indexed by the mean scanpath time, defined as the average duration of the seven successful scan paths per session. This reflects how long users take to complete each path in normal use; with higher values suggesting uncertainty or inefficient handling (Figure 2D). Consistency was indexed by the SD of scanpath time; a smaller SD indicates more uniform path durations and greater ease with the device. From baseline to follow-up, participants became both faster and more consistent, indicating growing comfort while maintaining good signal quality (p = 0.001; rank-biserial r = −0.64; Table 1; Figure 2D) These patterns are consistent with learning in which early users may need more time to scan well, followed by convergence toward an efficient, typical path duration as experience accrues.

To assess signal integrity independent of speed, we tracked two quality indicators. Lift on dark captures brief probe lifts during motion; lower values imply steadier contact and fewer motion artefacts (Figure 3A). Noise on dark reflects the noise level in the scan; lower values indicating cleaner ambient scan quality (Figure 3B). Across sessions, both indices remained stable, indicating that operators did not trade off signal quality while learning to work faster (Table 1). Specifically, lift on dark showed no reliable change (p = 0.466, d_z_ = −0.13; Figure 3A), and noise on dark was likewise unchanged (p = 0.524, *r*_*rb*_ = 0.13; Figure 3B). These findings support repeatable acquisition quality after brief practice.

**Figure 3.**
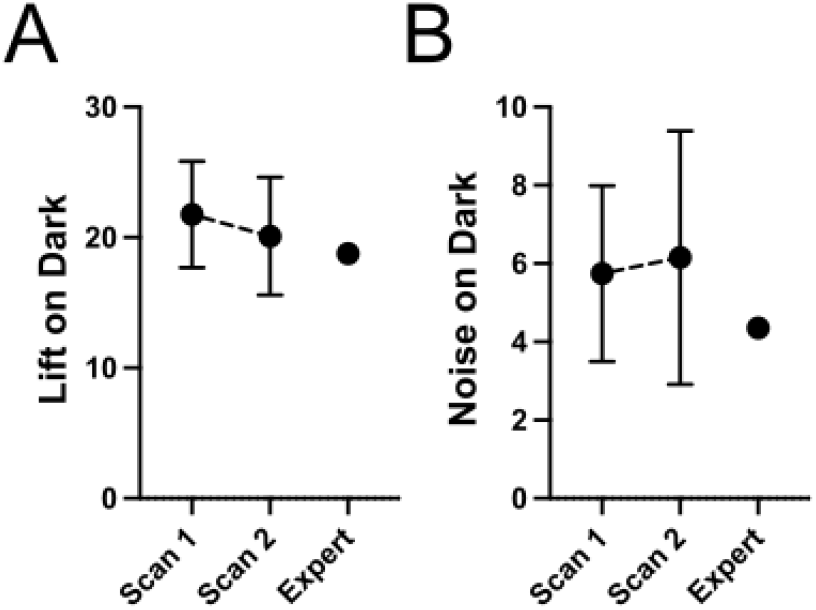
Scan-quality indicators remain stable across sessions. **(A)** Lift on dark and **(B)** Noise on dark are shown for novice operators after Scan 1 and Scan 2, with the expert reference plotted for context on the right of each panel. Points show group means with error bars, and lower values indicate better performance for both metrics.

After each scan, operators completed the 10-item System Usability Scale (SUS; 0-100, higher is better), covering ease of use, learnability, confidence, and perceived value. Usability ratings increased by about 7 points on the SUS from baseline to follow up with a moderate effect (p = 0.006, d_z_ = 0.52; Table 1; Figure 4), indicating a meaningful improvement in perceived ease of use after brief familiarization.

**Figure 4.**
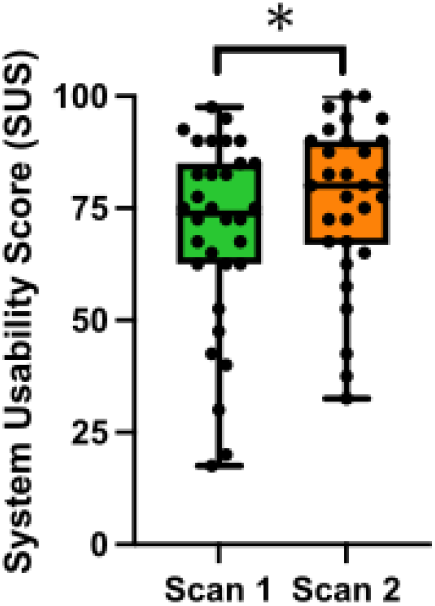
System Usability Scale (SUS) across two scans. Box-and-whisker plots display scores (0-100, higher indicates better usability) after Scan 1 (green) and Scan 2 (orange) with individual data overlaid.

## Discussion

This study demonstrates that non-specialist operators can achieve rapid proficiency in using th ArcheOptix NIRD^®^ device for head injury assessment. After one session, operators completed scans about 2.5 minutes faster, made fewer placement errors, and followed more consistent scanpaths. Quality metrics reflecting contact stability and ambient noise remained in a good range and did not change while users sped up, indicating that efficiency gains did not compromise signal quality. Perceived usability of the device also improved after a single usage, demonstrating a rapid uptake in familiarity. Together, these results suggest that reliable data can be acquired by new users with minimal practice, and that performance improves meaningfully from a single session. In practical terms, this supports the feasibility of deploying NIR screening with short, standardized training in prehospital and resource-limited settings where rapid triage is required.

CT remains the reference standard for diagnosing acute intracranial hemorrhage but is constrained by radiation, cost, and limited availability in pre-hospital, remote, and resource-limited environments.^3,6,9–11^ Portable NIR devices offer a non-ionizing, low-infrastructure screen that can support early healthcare decision making in pre-hospital, remote, and resource-limited environments.^4,5,12,17,18^ We used a simple task model in which operators followed illuminated paths at steady pressure and received built-in rescan cues when contact was lost. Our results address a practical bottleneck for such tools: whether non-experts can acquire adequate scans quickly. The observed learning curve and stable signal-quality suggest that the operational demands are compatible with frontline use after minimal training.

Participants rated the ArcheOptix device in the upper 70s on the System Usability Scale (SUS), which is a good rating above the average benchmark of ∼68.^19^ SUS is a validated, technology-agnostic measure with strong reliability and predictive value for acceptance.^15,20^ From baseline to follow-up, SUS increased by ∼7 points, a statistically meaningful rise consistent with improved confidence after minimal exposure. These gains reflect a noticeable improvement in user sentiment, and because usability perceptions rose alongside performance, it builds trust associated with higher likelihood of adoption in real-world settings.^15,20,21^ Easy to use devices are likely to become a regular part of triage, and provide a fast image among time-pressured clinicians.^4,22,23^

Several limitations shape how these results should be applied. Scans were performed on a healthy volunteer in a controlled setting, so distractions, hairstyles, or field constraints were not stress-tested. Retention was assessed over one day rather than weeks or months. Also, outcomes were device-specific; while concepts generalize, thresholds like the five-minute benchmark should be re-evaluated for other scanners and protocols. Finally, while the NIRD^®^ device is valuable for detecting the presence of intracranial bleeding, it does not provide the detail needed to localize a lesion, estimate volume, or guide surgical planning. Its strength is speed: it can flag a likely bleed within minutes at the point of care. Patients with positive or indeterminate screens should still undergo diagnostic imaging such as MRI to confirm the precise location. The practical value of this can be to reduce unnecessary transfers and airlifts and focus CT use on those most likely to benefit, provided performance is confirmed in real-world settings. These factors argue for a cautious targeted validation paradigm in the environments where the tool will be used. Our future work will address testing the device in realistic environments (hospital emergency departments) and include older operators.

Novices with minor training were able to operate a NIR head-injury scanner quickly and reliably, with faster scans, fewer rescans, stable signal quality, and higher usability ratings after first use. These results support NIR screening as a practical complement to CT that can help prioritize transport and focus limited imaging resources in emergency departments and resource-limited settings. The device is suited to rapid triage rather than definitive diagnosis, so positive or uncertain screens should still be followed by diagnostic imaging. Used appropriately, brief training with NIR screening can accelerate safe decision making when minutes matter.

## Data Availability

Data is available upon request

## Acknowledgements

This work was supported by NSERC for S.D., and SEAMO for D.J.C. Additionally for S.D., the research was undertaken thanks in part to funding from the Connected Minds Program, supported by Canada First Research Excellence Fund, Grant #CFREF-2022-00010. The funders had no role in study design, data collection and analysis, decision to publish, or preparation of the manuscript.

## Author Contributions

S.D., J.D.R. and D.J.C designed the research. S.D. and M.T.L. collected the data. S.D. and J.D.R. contributed experimental and analytic code. S.D. and J.D.R analyzed the data. S.D wrote the manuscript. All authors reviewed and approved the final manuscript.

## Data Availability Statement

Data is available upon request to the corresponding author.

## References

1. Dewan MC, Rattani A, Gupta S, et al. Estimating the global incidence of traumatic brain injury. J Neurosurg. 2019;130(4):1080–1097. doi:10.3171/2017.10.JNS17352

2. Maas AI, Stocchetti N, Bullock R. Moderate and severe traumatic brain injury in adults. The Lancet Neurology. 2008;7(8):728–741. doi:10.1016/S1474-4422(08)70164-9

3. Ibañez J, Arikan F, Pedraza S, et al. Reliability of clinical guidelines in the detection of patients at risk following mild head injury: results of a prospective study. J Neurosurg. 2004;100(5):825–834. doi:10.3171/jns.2004.100.5.0825

4. Zarei H, Zarrin A, Janmohamadi M, et al. Near Infrared Spectroscopy as a Diagnostic Tool for Screening of Intracranial Hematomas; A Systematic Review and Meta-Analysis. Arch Acad Emerg Med. 2024;13(1):e9. doi:10.22037/aaem.v13i1.2411

5. Shah J, Solanki S, Chandra A, Laljibhai Shivabhai M, Gwalani P. Evaluating performance of a near-infrared-spectroscopy-based and machine-learning-powered bio-optical sensitivity parameters in identifying intracranial hemorrhages in TBI across different age-groups. Brain Injury. 2024;38(14):1227–1235. doi:10.1080/02699052.2024.2381056

6. Brogan RJ, Kontojannis V, Garara B, Marcus HJ, Wilson MH. Near-infrared spectroscopy (NIRS) to detect traumatic intracranial haematoma: A systematic review and meta-analysis. Brain Inj. 2017;31(5):581–588. doi:10.1080/02699052.2017.1287956

7. Robertson CS, Zager EL, Narayan RK, et al. Clinical Evaluation of a Portable Near-Infrared Device for Detection of Traumatic Intracranial Hematomas. Journal of Neurotrauma. 2010;27(9):1597–1604. doi:10.1089/neu.2010.1340

8. Yuksen C, Sricharoen P, Puengsamran N, Saksobhavivat N, Sittichanbuncha Y, Sawanyawisuth K. Diagnostic properties of a portable near-infrared spectroscopy to detect intracranial hematoma in traumatic brain injury patients. Eur J Radiol Open. 2020;7:100246. doi:10.1016/j.ejro.2020.100246

9. Gramer R, Shlobin NA, Yang Z, Niedzwiecki D, Haglund MM, Fuller AT. Clinical Utility of Near-Infrared Device in Detecting Traumatic Intracranial Hemorrhage: A Pilot Study Toward Application as an Emergent Diagnostic Modality in a Low-Resource Setting. Journal of Neurotrauma. 2023;40(15-16):1596-1602. doi:10.1089/neu.2021.0342

10. Peters J, Van Wageningen B, Hoogerwerf N, Tan E. Near-Infrared Spectroscopy: A Promising Prehospital Tool for Management of Traumatic Brain Injury. Prehosp Disaster Med. 2017;32(4):414–418. doi:10.1017/S1049023X17006367

11. Leon-Carrion J, Dominguez-Roldan JM, Leon-Dominguez U, Murillo-Cabezas F. The Infrascanner, a handheld device for screening in situ for the presence of brain haematomas. Brain Injury. 2010;24(10):1193–1201. doi:10.3109/02699052.2010.506636

12. Viderman D, Ayapbergenov A, Abilman N, Abdildin YG. Near-infrared spectroscopy for intracranial hemorrhage detection in traumatic brain injury patients: A systematic review. Am J Emerg Med. 2021;50:758–764. doi:10.1016/j.ajem.2021.09.070

13. D’Amario S, Bougadis J, Coverdale NS, et al. A Handheld Near Infrared Scanner for the Detection of Acute Traumatic Intracranial Hemorrhage. Research Square. Preprint posted online December 12, 2025. doi:10.21203/rs.3.rs-8166633/v1

14. Riley JD, Amyot F, Pohida T, et al. A hematoma detector—a practical application of instrumental motion as signal in near infra-red imaging. Biomed Opt Express. 2012;3(1):192. doi:10.1364/BOE.3.000192

15. Brooke J. SUS - A quick and dirty usability scale. Usability Evaluation in Industry. Published online 1996.

16. R Core Team. R: A Language and Environment for Statistical Computing. R Foundation for Statistical Computing. Published online 2025. https://www.R-project.org/

17. Gibson A, Dehghani H. Diffuse optical imaging. Phil Trans R Soc A. 2009;367(1900):3055–3072. doi:10.1098/rsta.2009.0080

18. Hillman EMC, Amoozegar CB, Wang T, et al. In vivo optical imaging and dynamic contrast methods for biomedical research. Philosophical Transactions of the Royal Society A: Mathematical, Physical and Engineering Sciences. Published online November 28, 2011. doi:10.1098/rsta.2011.0264

19. Hyzy M, Bond R, Mulvenna M, et al. System Usability Scale Benchmarking for Digital Health Apps: Meta-analysis. JMIR mHealth and uHealth. 2022;10(8):e37290. doi:10.2196/37290

20. Bangor A, Kortum PT, Miller JT. An Empirical Evaluation of the System Usability Scale. International Journal of Human-Computer Interaction. 2008;24(6):574–594. doi:10.1080/10447310802205776

21. Sauro J, Lewis JR. When designing usability questionnaires, does it hurt to be positive? In: Proceedings of the SIGCHI Conference on Human Factors in Computing Systems. ACM; 2011:2215–2224. doi:10.1145/1978942.1979266

22. Healy J, Tzeng CFT, Wolfshohl J, et al. Point-of-Care Ultrasound in the Emergency Department: Training, Perceptions, Applications, and Barriers from Different Healthcare Professionals. J Acute Med. 2024;14(2):74–89. doi:10.6705/j.jacme.202406_14(2).0003

23. Nicola R, Dogra V. Ultrasound: the triage tool in the emergency department: using ultrasound first. Br J Radiol. 2016;89(1061):20150790. doi:10.1259/bjr.20150790

